# Blood group O and post-COVID-19 syndrome

**DOI:** 10.1101/2022.03.10.22272197

**Authors:** Sara Díaz-Salazar, Raquel Navas, Laura Sainz-Maza, Patricia Fierro, Meryam Maamar, Arancha Artime, Héctor Basterrechea, Benedetta Petitta, Carlota Lamadrid, Lucía Pedraja, Claudia Gándara-Samperio, Stefanie Pini, José Manuel Olmos, Carmen Ramos, Emilio Pariente, José Luis Hernández

**Affiliations:** Camargo Interior - Primary Health Care Center. Servicio Cántabro de Salud. Muriedas. Cantabria. Spain; Camargo Costa - Primary Health Care Center. Servicio Cántabro de Salud. Maliaño. Cantabria. Spain; Emergency Service. Osakidetza. Servicio Vasco de Salud. Bilbao. País Vasco. Spain; El Llano - Primary Health Care Center. SESPA- Servicio Asturiano de Salud. Gijón. Asturias. Spain; Hospital at Home Department. Hospital Universitario Marqués de Valdecilla. Santander. Cantabria. Spain; Depto. de Medicina y Psiquiatría. Universidad de Cantabria. Santander. Cantabria. Spain; Servicio de Medicina Interna. Hospital Universitario Marqués de Valdecilla. Instituto de Investigación Valdecilla (IDIVAL). Santander. Cantabria. Spain

**Keywords:** Blood group O, Post-COVID-19 syndrome, ABO blood group, COVID-19

## Abstract

**Objective:** The ABO blood group system modulates the inflammatory response and has been involved in COVID-19. O-group protects against SARS-CoV-2 infection, but there are no data regarding post-COVID-19 syndrome (PCS). Our aim was to assess this possible association.

**Subjects and methods:** Case-control study in a community setting, with subjects who had experienced mild COVID-19. Cases were PCS+, controls were PCS-, and the exposure variable, O-group. Epidemiological data (age, sex, BMI, smoking, comorbidities), laboratory parameters (inflammatory markers, IgG antibodies, blood type) and clinical data were collected. Composite inflammatory indices were developed. Multivariate analyses were performed.

**Results:** We analyzed 121 subjects (56.2% women), mean age 45.7 ± 16 years. Blood group frequencies were 43.3%, 7.7%, 5.7%, and 43.3% for A, B, AB and O, respectively. Thirty-six patients were PCS+. There were no significant differences between cases and controls. Compared to non-O, a higher prevalence of PCS (p=0.036), number of symptoms (p=0.017) and myalgia (p=0.030) were noted in O-group. Concerning inflammatory markers, PCS+ and PCS-showed no differences in A, B, and AB groups. In contrast, O-group PCS+ patients had significantly higher lymphocyte count, higher levels of fibrinogen and CRP, and higher percentages of 3 composite indices, than PCS-subjects. The O-group showed a 4-fold increased risk of PCS compared to non-O (adjusted OR=4.20 [95%CI, 1.2-14]; p=0.023).

**Conclusion:** An increased risk of PCS has shown to be associated with O-group, after controlling for confounders. In O-group subjects with PCS, slightly albeit significant, raised levels of fibrinogen, CRP, and lymphocyte count, have been demonstrated.

## INTRODUCTION

Blood groups A, B, AB, and O configure the ABO system, the most important blood group system in humans. It is located on chromosome 9 (band 9q34.2) and is composed of proteins and oligosaccharides expressed on the surface of red blood cells. Blood groups play a relevant role in cardiovascular and oncological diseases, thromboembolic and hemorrhagic processes, as well as infectious diseases (parasitic, bacterial, and viral), acting sometimes as receptors or co-receptors that facilitate viral particles from entering the cells [1]. People with blood group O have a lower risk of diabetes mellitus, arteriosclerosis, and ischemic heart disease [2], but a higher risk of tuberculosis, cholera, and Norovirus infections [3].

Since the beginning of the SARS-CoV-2 pandemic, a higher prevalence of COVID-19 in blood group A subjects and lower susceptibility to acquiring the infection in those with blood group O was noted [4,5]. This finding has been related to the anti-A antibodies presented in blood group O individuals. These antibodies bind themselves to the A-like antigens or to the spike protein expressed on the virus envelope, blocking its interaction with the ACE-2 receptor, which is an identified gateway for different coronaviruses to infect host cells [6].

Regarding clinical outcomes, blood group A has been associated with a higher risk of hospitalization, mechanical ventilation, and death [6,7]. These adverse outcomes have been related to higher levels of soluble circulating proteins linked to vascular adhesion in people with this group [8]. The studies are discordant concerning the clinical outcomes of group O patients [7,8]. Thus, some have observed a better prognosis [9], that has been related to lower circulating levels of factor VIII and von Willebrand factor -and a subsequently reduced risk of thrombotic phenomena-, and to lower angiotensin-converting enzyme (ACE) activity [10]. However, in other studies, group O has been related to serious outcomes [11-13].

After COVID-19, a variable percentage of patients, even those with mild initial forms of the disease, report prolonged and recurrent symptoms for weeks or months. These symptoms include fatigue -the most frequent-, myalgia, dyspnea, anosmia/ageusia, autonomic dysregulation manifested as orthostatic hypotension, tachycardia, thermoregulation or gastrointestinal disturbances, and cognition alterations, leading to a significant impact on the quality of life [14-16]. This condition, known as Long COVID or Post-COVID Syndrome (PCS) [17], has certain similarities with the chronic profile of other epidemic coronavirus infections, such as SARS and MERS [18].

A previous study by our group [19] has shown that in PCS, 3 months after the acute episode, there are slight but significant elevations of inflammatory markers, compared to controls who have had COVID-19 but not PCS, supporting the hypothesis of chronic low-grade inflammation underlying PCS pathogenesis [20,21]. On the other hand, the ABO system modulates endothelial function and influences the inflammatory response [8,22], although the mechanisms are not fully understood [8].

Considering the associations of the ABO system with the inflammatory response, the evidence of the relationship between the ABO system and COVID-19, and PCS as an expression of low-grade inflammation, it is worth asking whether the protective effect of group O against infection extends to the post-acute phase. Taking into account the above considerations, we aimed to assess whether blood group O is related to PCS after mild COVID-19.

## SUBJECTS AND METHODS

### Design

A case-control study including subjects who had suffered from mild COVID-19 in a community setting was designed. Cases were PCS+ patients, controls were PCS-subjects and the exposure variable was blood group O. The study was conducted on the general population of a semi-urban area attended by a Primary Care center in Northern Spain. The general design of the study is shown in Figure 1.

**Figure 1:**
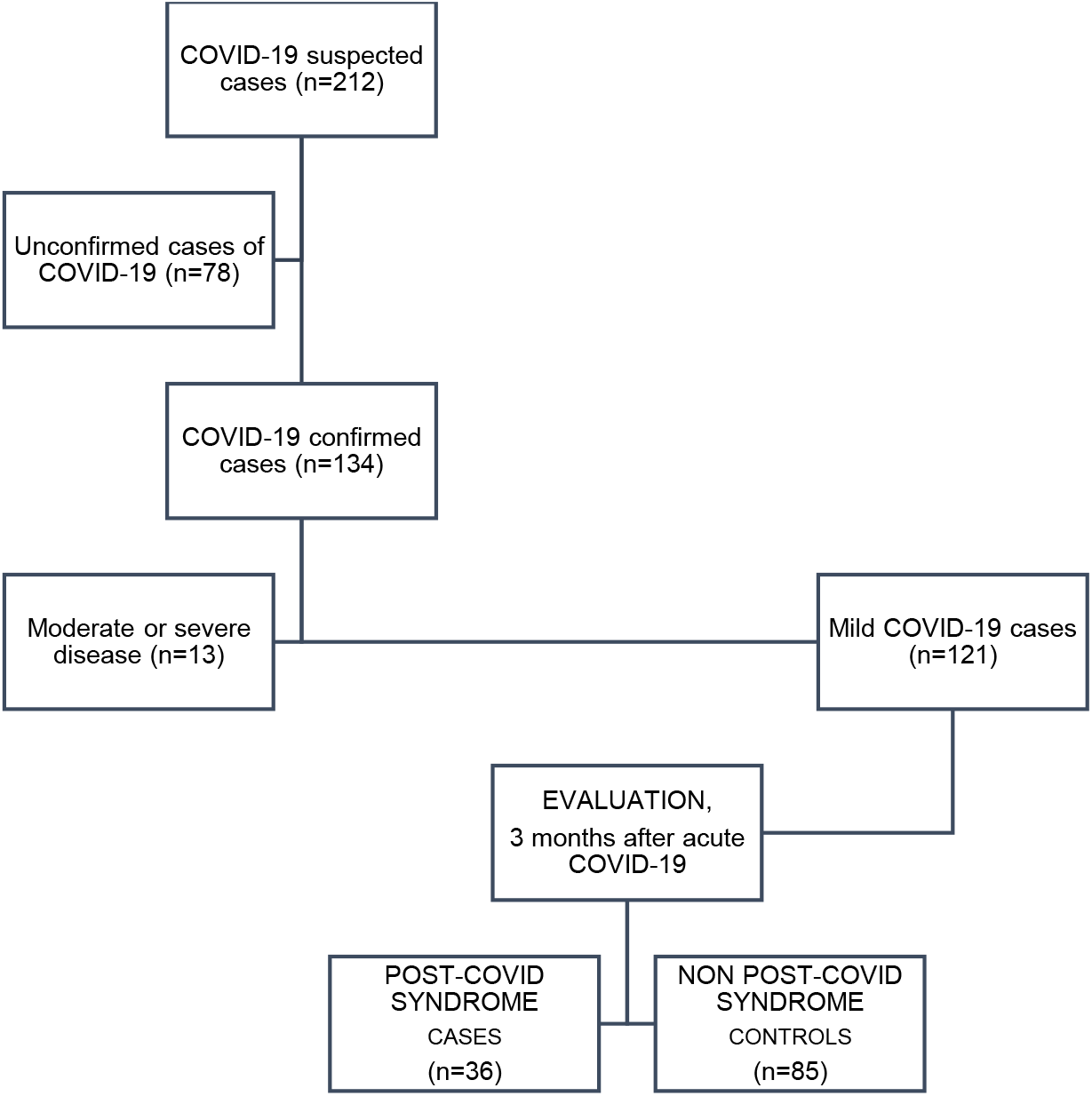
Flow chart and study design

### Participants

Details of participants’ enrollment have been previously published [19]. Patients suffering from COVID-19 between April and September 2020 were selected. All the cases were exclusively followed in the Primary Care setting and none of them had been vaccinated against SARS-CoV-2 at the moment of inclusion. SARS-CoV-2 infection was confirmed by a positive real-time reverse transcription-polymerase chain reaction (RT-PCR) test or by the presence of anti-SARS-CoV-2 IgG, three months after COVID-19. A second inclusion criterion was a mild course of infection, according to the WHO definition [23] and characterized by fever, malaise, cough, upper respiratory symptoms, and/or less common COVID-19 manifestations, in the absence of dyspnea. The choice of outpatients with mild COVID-19 reasonably ruled out post-acute symptoms due to previous comorbidities or organ sequelae [19]. No exclusion criteria were considered.

### Data collection

Three months after the acute episode (median=115 days), epidemiological variables (sex, age, body mass index -BMI, measured in kg/m^2^-, smoking habit, medical history, and Charlson’s comorbidity index), clinical data (specific symptoms and number of symptoms -as a variable assimilated to ‘intensity’ of the PCS [24]-) and laboratory test (inflammation markers and anti-SARS-CoV-2 IgG antibodies) were collected. One year later, participants were interviewed once again, and their blood group and Rh factor were obtained.

The interviews were conducted by physicians from the research team, using a structured questionnaire. PCS diagnosis was established when the National Institute for Care and Excellence (NICE) criteria was met: signs and symptoms that develop during or after an infection consistent with COVID-19, that last more than 12 weeks, and that are unexplained by an alternative diagnosis [17].

### Serum inflammatory markers

Serum C-reactive protein (CRP), ferritin, lactate dehydrogenase (LDH), fibrinogen and D-dimer levels, as well as neutrophil and lymphocyte counts, and neutrophil/lymphocyte ratio (NLR) were analyzed. CRP was measured in mg/dL, and the detection limit was 0.4 mg/dL. Low-grade inflammation has been defined by a serum CRP level >0.3 mg/dL and <1.0 mg/dL [25]. Normal ranges for LDH (U/L), ferritin (ng/mL), fibrinogen (mg/dL), D-dimer (ng/mL), neutrophil count, and lymphocyte count were 120-246, 22-322, 180 -500, 0-500, 1.4-7.5 (x10^3^/µL), and 1.2-5 (x10^3^/µL), respectively.

Blood samples were obtained from an antecubital vein using the standard venipuncture procedure in the morning after a 12-hour fast. LDH and ferritin were analyzed by spectrophotometric assay on an Atellica CH analyzer (Siemens Healthcare Diagnostics Inc, Tarrytown, NY, USA). CRP was quantified by immunonephelometric assay on an Atellica CH analyzer (Siemens Healthcare Diagnostics Inc, Tarrytown, NY, USA). Hematologic cell counts were analyzed on a DXH900 (Beckman Coulter), and fibrinogen and D-dimer on an ACL TOP 750 (Werfen). Anti-N IgG was obtained by chemiluminescence (QLIA), expressing the result as positive or negative.

### Blood group samples

The blood group was determined in capillary blood when it was not available in the clinical chart. To do this, a blood sample was taken by puncturing the pad of the finger with a lancet. The visualization or not of the agglutination reaction allowed the identification of the blood group when confronting the sample with reagents of known specificity (Alden Reagents, Laboquimia, Spain)

### Statistical analysis

After assessing normality with the Shapiro-Wilk test, quantitative variables were expressed as mean ± standard deviation (SD) or as median [interquartile range (IQR)]. Some variables have been expressed as dichotomous using the median or tertiles. Student’s t-test and ANOVA were used as parametric tests and the median, Mann-Whitney U and Kruskal-Wallis tests, as non-parametric procedures. Categorical variables have been expressed as numbers and percentages and the chi-square tests were used for their comparison. Correlation analyses have been performed using Spearman’s Rho and the Phi coefficient for categorical variables. The strength of an association has been expressed as an odds ratio (OR) with its corresponding 95% confidence interval (CI). Given the small number of subjects in blood groups B and AB, they were classified as group O/non-O, group A/non-A, group B/non-B, and group AB/non-AB.

After selecting the serum inflammatory markers that showed the highest correlations with PCS, the upper ranges of their distributions were combined to develop the composite inflammatory indices, which were designated correlatively as C1-C5 (Table 1).

**Table 1:** Composite indices of inflammation

| COMPOSITE INDEX | DEFINITION |
| --- | --- |
| C1 | [Neutrophil count $\geq 3.10^*$ ($\times 10^3/\mu\text{L}$)] or [NL Ratio $\geq 1.86^{**}$] |
| C2 | [CRP $> 0.3$ and $< 1.0$ mg/dL] or [Fibrinogen $\geq 421^{**}$ mg/dL] |
| C3 | [Neutrophil count $\geq 3.10^*$ ($\times 10^3/\mu\text{L}$)] or [Fibrinogen $\geq 421^{**}$ mg/dL] |
| C4 | [NL Ratio $\geq 1.86^{**}$] or [Fibrinogen $\geq 421^{**}$ mg/dL] |
| C5 | [CRP $> 0.3$ and $< 1.0$ mg/dL] or [Neutrophil count $\geq 3.40^{**}$ ($\times 10^3/\mu\text{L}$)] |

CRP: C-reactive protein; NL Ratio: Neutrophil / Lymphocyte Ratio
\*Values above the median; \*\*Values in the third tertile

Logistic regression was performed to determine the relationship between group O and PCS, after controlling for confounding variables selected according to the literature and the principle of parsimony. The model was validated using the determination coefficient R^2^ and the area under the ROC curve (AUC).

IBM SPSS 28.0 statistical package (Armonk, NY: IBM Corp) was used to perform the analyses. A *p*-value <0.05 was considered significant in all calculations.

### Ethical aspects

The postulates of the Declaration of Helsinki were fulfilled. The study was approved by the Clinical Research Ethics Committee of Cantabria (Internal Code 2021.102).

## RESULTS

### Descriptive analysis

One-hundred and thirty-four patients with confirmed COVID-19 were included, and 13 were discarded due to moderate or severe disease (Figure 1). Thus, we finally included 121 subjects with mild COVID-19, 68 of them (56.2%) were women. The mean age was 45.7±16 years (range, 18-88 years). The frequencies of the blood groups were 43.3% (group A), 7.7% (group B), 5.7% (group AB) and 43.3% (group O). Tertiles of age distribution were <41, 41-53, >53 years and BMI, <23.8, 23.8-27.6, >27.6 Kg/m^2^. Thirty-six subjects, 29.7% of the sample, were classified as PCS+; 25 were women (35.8%) and 11 men (20.8%) (p=0.07). The remaining 85 were classified as PCS-. The most frequent PCS symptoms were fatigue (42.8%), anosmia (40%), ageusia (22.8%), dyspnea (17.1%), myalgia (11.4%) and palpitations (11.4%).

Table 2 shows the baseline characteristics of the PCS+ and PCS-participants. There were no significant differences between both groups concerning BMI, tobacco use, or previous comorbidities.

**Table 2:** Clinical characteristics of cases and controls

|  | **CASES**<br>**(PCS+)**<br>**(n=36)** | **CONTROLS**<br>**(PCS-)**<br>**(n=85)** | **P** |
| --- | --- | --- | --- |
| Females; n(%) | 25 (35.8) | 43 (64.2) | 0.07 |
| Males; n(%) | 11 (20.8) | 42 (79.2) |  |
| Age (years); mean ± SD | 46.7 ± 14 | 45.1 ± 17 | 0.62 |
| Age: tertile 1; n(%) | 9 (25.7) | 32 (38.6) | 0.48 |
| Age: tertile 2; n(%) | 16 (45.7) | 26 (31.3) |  |
| Age: tertile 3; n(%) | 10 (28.6) | 25 (30.1) |  |
| BMI; mean ± SD | 24.7 ± 4 | 25.6 ± 3 | 0.28 |
| Obesity; n(%) | 3 (8.6) | 12 (15.2) | 0.33 |
| Tobacco; n(%) | 11 (31.4) | 32 (39) | 0.43 |
| Charlson comorbidity index; mean ± SD | 0.20 ± 0.4 | 0.48 ± 0.8 | 0.08* |
| CCI=0; n(%) | 28 (80) | 56 (68.3) | 0.15 |
| CCI=1; n(%) | 7(20) | 24 (29.3) |  |
| CCI=2+; n(%) | - | 2 (2.4) |  |
| Blood group A; n(%) | 11 (33.3) | 33 (47.1) | 0.09 |
| Blood group B; n(%) | 2 (6.1) | 6 (8.6) |  |
| Blood group AB; n(%) | 1 (3) | 5 (7.1) |  |
| Blood group O; n(%) | 19 (57.6) | 26 (37.1) |  |
| Rh+; n(%) | 25 (75.8) | 53 (75.7) | 0.95 |
| Rh-; n(%) | 8 (24.2) | 16 (24.3) |  |
| Immunosuppression; n(%) | 1 (2.9) | 2 (2.4) | 0.89 |
| Chronic kidney disease; n(%) | - | 2 (2.4) | 0.95 |
| Cerebrovascular disease; n(%) | - | 1 (1.2) | 0.98 |
| Hypertension; n(%) | 4 (11.4) | 17 (20.7) | 0.23 |
| Diabetes mellitus; n(%) | 1 (2.9) | 5 (6.1) | 0.60 |
| Dyslipidemia; n(%) | 12 (34.3) | 26 (31.7) | 0.78 |
| Ischemic heart disease; n(%) | 2 (5.7) | 4 (4.9) | 0.85 |
| Asthma; n(%) | 5 (14.3) | 6 (7.3) | 0.30 |

PCS+: With post-COVID syndrome; PCS-: Without post-COVID syndrome. SD: standard deviation; BMI: Body mass index \*Mann-Whitney U-test

### Post-COVID syndrome

The prevalence of PCS according to the blood group was: 25% (group A) and 37.3% (non-A) (p=0.18); 25% (group B) and 32.6% (non-B) (p=0.65); 16.7% (AB group) and 33% (non-AB) (p=0.40); 43.2% (O group) 23.7% (non-O) (p=0.036); 32.1% (Rh+) and 32% (Rh-) (p=0.99).

The mean number of symptoms of PCS was 0.44±0.9 (group A) and 0.71±0.9 (non-A) (p=0.17); 0.50±0.9 (group B) and 0.60±1 (non-B) (p=0.77); 0.33±0.8 (AB group) and 0.61±1 (non-AB) (p=0.50); 0.82±1 (O group) and 0.43±0.9 (non-O) (p=0.017); 0.59±0.9 (Rh+) and 0.60±1 (Rh-) (p=0.98). The symptoms analyzed were anosmia, dyspnea, palpitations, asthenia, telogen effluvium, ageusia, headache, leukonychia, myalgia, concentration difficulties, rhinitis, and cough. They were evaluated according to their presence in O and non-O subjects, with myalgia presenting a significant difference between both groups (Table 3). When analyzing these patients, it has been observed that both group O subjects and patients with myalgia have presented a high frequency of positives in the C2 composite index (64.7% of group O vs 22.7% of non-O, p=0.007; 100% of subjects with myalgia vs 46.2% of subjects without myalgia, p=0.044). Regarding biomarkers, fibrinogen levels in patients with and without myalgia were 510±82 mg/dL and 394.6±87, respectively (p=0.013).

**Table 3:** Clinical characteristics and variables related to PCS in group O and non-O subjects

|  | GROUP O<br>(n=44) | NON-O<br>(n=60) | p* |
| --- | --- | --- | --- |
| Females; n(%) | 26 (40.6) | 38 (59.4) | 0.66 |
| Males; n(%) | 18 (45) | 22 (55) |  |
| Age (years); mean ± SD | 46.9 ± 16 | 47.3 ± 16 | 0.90 |
| Age: tertile 1; n(%) | 15 (46.9) | 17 (53.1) | 0.79 |
| Age: tertile 2; n(%) | 14 (38.9) | 22 (61.1) |  |
| Age: tertile 3; n(%) | 14 (41.2) | 20 (58.8) |  |
| BMI; mean ± SD | 25.1 ± 3 | 25.5 ± 4 | 0.66 |
| Obesity; n(%) | 5 (11.6) | 8 (14.5) | 0.67 |
| Tobacco; n(%) | 12 (27.9) | 26 (44.8) | 0.08 |
| Charlson comorbidity index; mean ± SD | 0.42 ± 0.8 | 0.43 ± 0.9 | 0.92* |
| Post-COVID-19 syndrome (+); n(%) | 19 (43.2) | 14 (23.7) | 0.036 |
| Number of symptoms; mean ± SD | 0.82 ± 1 | 0.43 ± 0.9 | 0.017* |
| Anosmia; n(%) | 5 (11.4) | 7 (11.7) | 0.96 |
| Dyspnea; n(%) | 2 (4.8) | 4 (6.7) | 0.64 |
| Palpitations; n(%) | 1 (2.3) | 3 (5) | 0.47 |
| Fatigue; n(%) | 9 (20.5) | 6 (10) | 0.13 |
| Telogen effluvium; n(%) | 2 (4.5) | 1 (1.7) | 0.57 |
| Ageusia; n(%) | 5 (11.4) | 3 (5) | 0.27 |
| Headache; n(%) | 2 (4.5) | 1 (1.7) | 0.57 |
| Leukonychia; n(%) | 1 (2.3) | - | 0.42 |
| Myalgia; n(%) | 4 (9.1) | - | 0.030 |
| Concentration difficulties; n(%) | 3 (6.8) | - | 0.07 |
| Rhinitis; n(%) | 1 (2.3) | - | 0.42 |
| Cough; n(%) | 1 (2.3) | - | 0.42 |
| Anti-SARS-CoV-2 IgG(+); n(%) | 34 (77.3) | 54 (90) | 0.07 |

*\* Mann-Whitney U-test*

Group O, fatigue, and serum CRP have shown to be related. In group O individuals, fatigue and raised serum CRP showed a positive correlation (Phi=0.372; p=0.022), while in non-O, fatigue and serum CRP were not correlated (Phi=0.03; p=0.80). In group A patients with fatigue, raised CRP was registered in 20%. By contrast, in group O patients with fatigue, the percentage was 37.5% (p=0.021)

### Anti-SARS-CoV-2 IgG antibodies

Ninety-eight persons (81%) had anti-SARS-CoV-2 IgG. IgG+ subjects had a mean age of 47.3±15 years, and IgG-subjects, 38.8±18 (p=0.030). IgG+ subjects presented 0.61±1 symptoms compared to 0.17±0.4 symptoms in IgG-subjects (p=0.044). No differences were observed between IgG+ and IgG-subjects regarding sex distribution, Charlson index, tobacco use, or serum inflammatory marker levels. Some 91.4% of subjects with PCS had IgG antibodies, compared to 76.5% of subjects without PCS (p=0.058).

The prevalence of anti-N IgG+ was 77.3% in the blood group O and 90% in the non-O group (p=0.076). In the remaining groups, these figures were 88.9% (group A), 100% (group B), 83.3% (group AB), 84.8% (Rh+) and 84.0% (Rh-).

### Inflammatory markers and composite indices

Table 4 shows correlation analyses between the 8 inflammatory markers and the number of symptoms of PCS. In blood group O, significant correlations were observed with lymphocyte count (r=0.334; p=0.025), fibrinogen level (r=0.404; p=0.009) and serum CRP (Phi=0.465; p=0.013). Rh+ people showed significant direct correlations of the number of symptoms with the count of neutrophils (Rho=0.247; p=0.029) and lymphocytes (Rho=0.253; p=0.026).

**Table 4:** Correlation analyses between inflammatory markers and the number of symptoms in PCS

|  | GROUP A |  | GROUP B |  | GROUP AB |  | GROUP O |  |
| --- | --- | --- | --- | --- | --- | --- | --- | --- |
|  | *r* | *p* | *r* | *p* | *r* | *p* | *R* | *p* |
| Neutrophil count (x10<sup>3</sup>/μL) | -0.050 | 0.75 | 0.632 | 0.12 | -0.266 | 0.61 | 0.194 | 0.20 |
| Lymphocyte count (x10<sup>3</sup>/μL) | -0.025 | 0.87 | 0.001 | 0.99 | 0.664 | 0.15 | 0.343 | **0.023** |
| Neutrophil/Lymphocyte Ratio | 0.004 | 0.97 | 0.316 | 0.49 | -0.665 | 0.15 | 0.126 | 0.41 |
| Ferritin (ng/mL) | -0.058 | 0.71 | -0.239 | 0.60 | -0.665 | 0.15 | -0.059 | 0.71 |
| Lactate dehydrogenase (UI/L) | 0.251 | 0.10 | -0.474 | 0.28 | -0.393 | 0.44 | -0.241 | 0.13 |
| Fibrinogen (mg/dL) | 0.067 | 0.68 | 0.158 | 0.73 | -0.393 | 0.44 | 0.384 | **0.014** |
| D-dimer (ng/mL) | -0.082 | 0.63 | 0.001 | 0.98 | 0.131 | 0.80 | 0.266 | 0.10 |
| CRP (mg/dL) ‡ | 0.068 | 0.91 | - | - | - | - | 0.462 | **0.017** |

CRP: C-reactive protein
‡ Correlation analysis between plasma CRP levels (<0.4 mg/dL and 0.4-0.9 mg/dL) and the number of symptoms of the PCS (expressed as 0, 1, 2 or more) was assessed with Phi coefficient.

The performance of biomarkers and composite indices of inflammation (C1 to C5), concerning PCS in each blood group, was also analyzed. Groups A, B, AB, and Rh-subjects did not present any significant difference between PCS+ and PCS-. Conversely, significant differences were observed in blood group O, in which PCS+ patients had a lymphocyte count, serum CRP and fibrinogen levels, and percentages of positivity in C2, C3, and C4 indices, significantly higher than PCS-subjects (Table 5 and Figure 2). Compared to the Rh+ subjects without PCS, those Rh+ with PCS had higher percentage of serum CRP in the low-grade inflammation range (28.6% vs. 8.5%; p=0.031), and higher percentage of subjects meeting the C1 (76 % vs. 50%; p=0.030), C3 (82.6% vs. 54%; p=0.019) and C4 criteria (69.6 vs. 44%; p=0.042).

**Table 5:** Inflammation markers and composite indices of inflammation in PCS, in relation to ABO system groups.

|  | GROUP A<br>(n=42) |  |  | GROUP B<br>(n=7) |  |  | GROUP AB<br>(n=6) |  |  | GROUP O<br>(n=45) |  |  |
| --- | --- | --- | --- | --- | --- | --- | --- | --- | --- | --- | --- | --- |
|  | PCS+<br>(n=11) | PCS-<br>(n=31) | p | PCS+<br>(n=2) | PCS-<br>(n=5) | p | PCS+<br>(n=1) | PCS-<br>(n=5) | p | PCS+<br>(n=19) | PCS-<br>(n=26) | p |
| Neutrophil count (x10<sup>9</sup>/μL) | 3.1±1.1 | 3.1±1.1 | 0.99 | 3.9±0.6 | 2.9±0.6 | 0.13 | 1.8 | 2.2±1.2 | 0.78 | 3.2±1.2 | 3±1 | 0.66 |
| Lymphocyte count (x10<sup>9</sup>/μL) | 1.8±0.5 | 1.9±0.7 | 0.60 | 2.1±0.2 | 2.1±0.7 | 0.98 | 2.2 | 1.5±0.2 | 0.06 | 2.1±0.8 | 1.5±1 | **0.033** |
| Neutrophil/Lymphocyte Ratio | 1.7±0.7 | 1.6±0.5 | 0.67 | 1.8±0.4 | 1.5±0.7 | 0.58 | 0.8 | 1.9±0.6 | 0.18 | 1.7±0.5 | 1.6±0.7 | 0.76 |
| Ferritin (ng/mL) | 46 [122] | 50.5 [99] | 0.64 | 50.5 | 81.5 [81] | 0.57 | - | 185 [337] | 0.33 | 67 [124] | 57 [104] | 0.98 |
| Lactate dehydrogenase (U/L) | 189±2.3 | 169±29 | 0.05 | 144±66 | 187±20 | 0.19 | 157 | 193 [45] | 0.51 | 172.4±27 | 190±35 | 0.08 |
| Fibrinogen (mg/dL) | 410±77 | 406±11 | 0.91 | 406±66 | 419±74 | 0.83 | 310 | 383±69 | 0.38 | 439±24 | 383±71 | **0.024** |
| D-dimer (ng/mL) | 199 [478] | 286 [230] | 0.61 | 308 [-] | 399 [233] | 0.99 | - | 242 [462] | 0.99 | 319 [407] | 201 [356] | 0.44 |
| CRP>0.3 and <1.0 mg/dL; n(%) | 2 (28.6) | 5 (71.4) | 0.95 | - | - | - | - | - | - | 5 (31) | - | **0.008** |
| C1 index (+); n(%) | 8 (72.3) | 16 (51.6) | 0.30 | 2 (100) | 2 (40) | 0.42 | - | 3 (60) | 0.95 | 15 (78.9) | 14 (53.8) | 0.08 |
| C2 index (+); n(%) | 3 (37.5) | 9 (30) | 0.68 | 1 (50) | 3 (60) | 0.95 | - | 1 (20) | 0.99 | 11 (64.7) | 5 (22.7) | **0.007** |
| C3 index (+); n(%) | 6 (66.7) | 16 (53.3) | 0.70 | 2 (100) | 4 (80) | 0.98 | - | 2 (40) | 0.98 | 17 (89.5) | 13 (52) | **0.008** |
| C4 index (+); n(%) | 6 (66.7) | 15 (50) | 0.46 | 1 (50) | 3 (60) | 0.96 | - | 4 (80) | 0.33 | 14 (77.8) | 8 (32) | **0.003** |
| C5 index (+); n(%) | 3 (33.3) | 13 (44.8) | 0.70 | 2 (100) | 1 (25) | 0.48 | - | 1 (20) | 0.99 | 10 (55.6) | 7 (28) | 0.06 |

PCS+: With post-COVID-19 syndrome; PCS-: Without post-COVID-19 syndrome; CRP: C-reactive protein Contrast test for quantitative variables: Student's t-test (except for ferritin and D-dimer: Mann-Whitney U).

**Figure 2:**
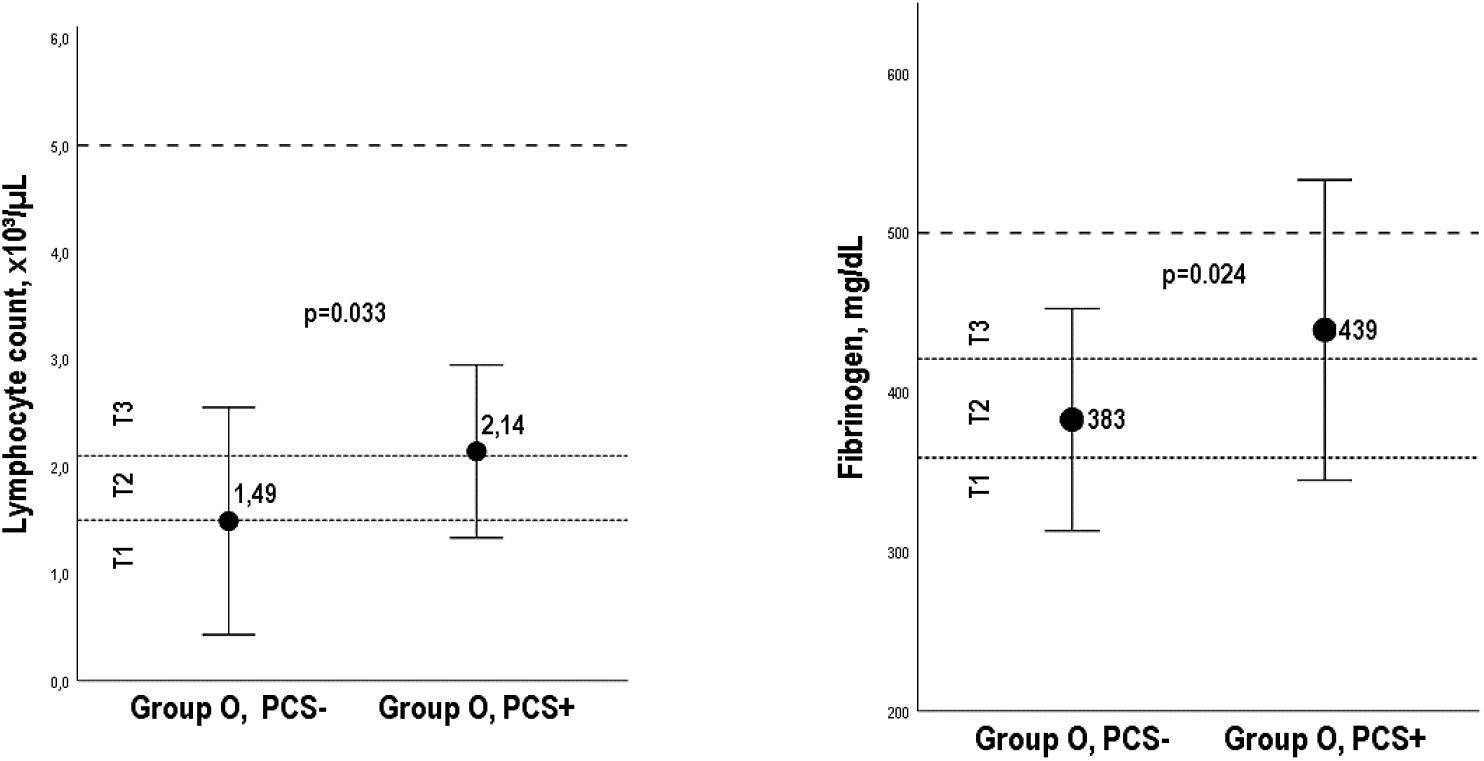
Lymphocyte count and plasma fibrinogen in group O patients with and without PCS Mean plot with error bars representing ± 1 standard deviation. PCS-: Without post-COVID syndrome; PCS+: With post-COVID syndrome. T1, T2, T3: Tertiles (Fibrinogen: <359, 359-421, >421 mg/dL; lymphocyte count: <1.5, 1.5-2.1, >2.1 x10^3^μL). Dashed lines at 5.0 x10^3^μL and at 500 mg/dL represent the upper limit of the normal range.

### Multivariate analysis

Table 6 shows the crude and adjusted OR (aOR) associated with the different blood groups and the Rh factor. Although in the crude analysis, groups B, AB, and Rh-showed significant ORs, the association was canceled when adjusting for age and sex. However, the OR for the blood group O remains significant. Additional adjustments for different clinical and laboratory confounders did not virtually modify this result. Blood group O subjects had a 4-fold increased risk of PCS, compared to the non-O group. Specifically, OR was 4.20 [95% CI, 1.2-14] (p=0.023), after adjusting for age, sex, BMI, tobacco use, comorbidity, Rh factor, neutrophil count, lymphocyte count, serum fibrinogen and CRP levels, hypertension, DM, and DLP. Nagelkerke’s R^2^ value was 0.46, and the AUC was 0.807 [95% CI 0.70-0.90] (p=0.0001) (Figure 3).

**Table 6:** Odds ratios associated with ABO groups and Rh factor, with PCS as outcome variable

|  | Crude analysis |  |  | Sex- and age-adjusted |  |  | Adjusted by sex, age, and additional adjustments* |  |  |
| --- | --- | --- | --- | --- | --- | --- | --- | --- | --- |
|  | OR | CI 95% | *p* | OR | CI 95% | *p* | OR | CI 95% | *p* |
| **Group A** | 1.68 | 0.9 – 2.8 | 0.053 | - | - | - | - | - | - |
| **Group B** | 2.06 | 1.3 – 3.1 | 0.001 | 0.79 | 0.2 – 2.8 | 0.72 | - | - | - |
| **Group AB** | 2.03 | 1.3 – 3.1 | 0.001 | 0.84 | 0.2 – 2.9 | 0.78 | - | - | - |
| **Group O** | 3.21 | 1.7 – 5.8 | 0.0001 | 2.49 | 1.08 – 5.7 | 0.031 | 4.20 | 1.2 – 14 | 0.023 |
| **Rh+** | 2.12 | 0.9 – 4.9 | 0.079 | - | - | - | - | - | - |
| **Rh-** | 2.12 | 1.3 – 3.4 | 0.002 | 1.01 | 0.3 – 2.5 | 0.97 | - | - | - |

*CI: Confidence interval*
\* *Body mass index, tobacco use, Charlson index, Rh factor, neutrophil count, lymphocyte count, fibrinogen level, serum C-reactive protein, hypertension, diabetes mellitus and dyslipidemia.*

**Figure 3:**
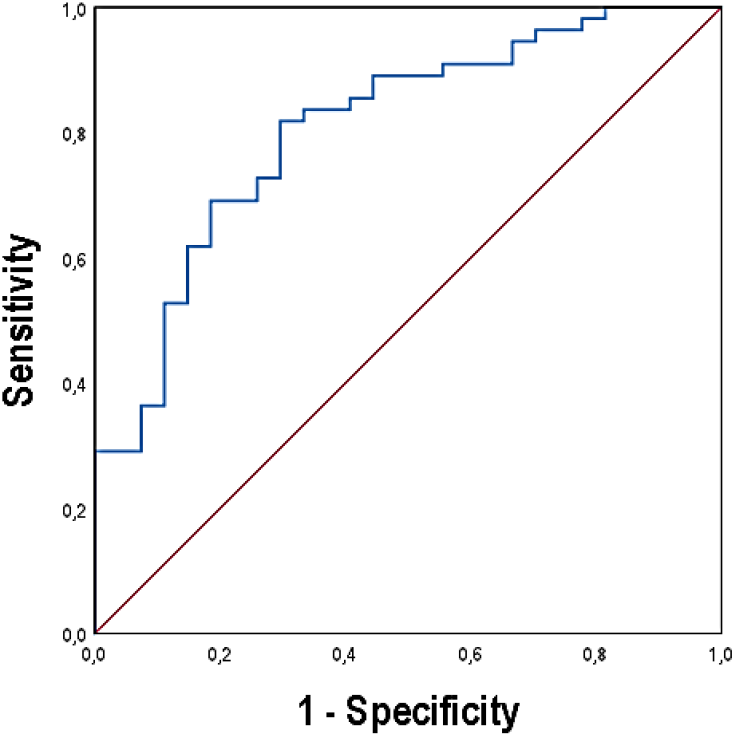
ROC curve associated with the logistic regression model, with group O as the independent variable and PCS as the outcome variable. *Area under the curve (AUC) = 0*.*807 (95% CI 0*.*70-0*.*90); p=0*.*0001*

## DISCUSSION

To our knowledge, this is the first study that found an independent association between blood group O and PCS. The relationships of blood group O with the PCS, inflammatory markers, fatigue, and myalgia are discussed below.

### Blood group O, PCS, and inflammation

Compared to non-O subjects, group O had a significantly higher prevalence of PCS and a higher number of PCS symptoms. In these patients (group O and PCS+), the mild elevated levels of biomarkers and the higher percentage of individuals meeting the criteria of the composite indices C2, C3 and C4, would point to a low-grade inflammation. These results are in line with the current knowledge since there is increasing evidence of an excessive and dysregulated inflammatory/immune activation underlying PCS pathogenesis [14,26]. However, the question of why PCS has been related to inflammatory markers in subjects with blood group O, and not in other groups of the ABO system, does not have a simple answer.

A genomic study by Naitza *et al*. has linked blood group O with increased levels of interleukin (IL) 6 (IL-6) [27]. IL-6 is a multifunctional proinflammatory cytokine involved in a wide range of immunomodulatory processes. It also has a relevant role in acute and chronic inflammation [28], activating the adhesion of lymphocytes and endothelial cells and influencing the magnitude of the inflammatory response [8]. In addition, it stimulates the synthesis of fibrinogen and CRP and regulates the recruitment of neutrophils and their subsequent controlled elimination [28-30]. Once the acute phase of COVID-19 is finished, IL-6 levels remain elevated in a variable percentage of patients, between 3.9% and 32.2% according to a recent review [14].

Based on what is known, it can be speculated that that some group O subjects, perhaps with basal raised levels of IL-6, after the acute COVID-19, could have maintained persistently high levels of IL-6. This could have stimulated the synthesis of CRP, fibrinogen, and other acute phase-reaction proteins, and promoted a prolonged inflammatory response, expressed clinically as a PCS.

In our opinion, this sequence could explain, at least in part, (i) the higher prevalence of PCS and myalgia, and the higher number of symptoms observed in group O compared to non-O, (ii) within the group O, the significant correlations between the number of symptoms and CRP, lymphocyte count and fibrinogen level, (iii) the significant percentage of C2 composite index (defined by high ranges of CRP or fibrinogen) presented by subjects with myalgia and by group O subjects, (iv) the differences between PCS+ and PCS-subjects observed in group O, but not in the other blood types, and (v) the group O as a consistent risk factor for PCS.

### Blood group O, fatigue, and CRP in range of low-grade inflammation

The persistent fatigue was the most self-reported symptom by participants, in line with the published papers [31]. A systematic review has shown that the pooled proportion of subjects with fatigue 12 or more weeks after the COVID-19 diagnosis was 0.32 (95% CI 0.27-0.37; p<0.001; n=25268) [14].

The exact mechanisms of chronic fatigue remain unraveled, but there is consensus in accepting 4 models: the oxidative stress and mitochondrial dysfunction, the hypothalamus-pituitary-adrenal axis, genetics and the inflammation [32]. Inflammation causes fatigue, and higher levels of circulating pro-inflammatory cytokines (tumor necrosis factor (TNF)-α, IL-1β, IL-6) and CRP than controls, have been noted in chronic fatigue [33,34]. In the same way, post-COVID fatigue has shown to be related to inflammatory markers [35-37]. The relation between fatigue and group O in COVID-19 has also been reported [38].

The results corroborate these findings. We found a significant association between fatigue, CRP in range of low-grade inflammation, and group O, not observed in the other blood groups.

Of note, in our previous work, gender showed to be a relevant variable in the relationship between fatigue and CRP, being significant only in men [19]. The correlation between fatigue and high CRP has been positive and quite similar in both subsamples (Phi=0.383; p=0.012, in men, and Phi=0.372; p=0.022 in group O).

### Blood group O, myalgia, and the possible role of fibrinogen

Subjects with blood group O had a significant association with post-COVID myalgia, a relationship that remained significant after adjusting for gender. A potential role of fibrinogen can be suggested as a pathophysiological marker. Thus, as previously said, blood group O has been associated with raised levels of IL-6, which is an important regulator of fibrinogen synthesis [39]. Secondly, apart from being involved in atherogenesis, thrombogenesis, and inflammation [40], fibrinogen is closely related to chronic pain [41,42]. In this context, fibrinogen has shown high sensitivity to changes in the activity of polymyalgia rheumatica and, therefore, it is used as a marker of the disease [43]. Along with other proteins, it has also been involved in the intensity of pain in fibromyalgia [44]. Finally, a proteomics study [45] has shown that at least 2 months after acute COVID-19, raised levels of inflammatory molecules, such as α2-antiplasmin, chains of fibrinogen, and serum amyloid A (SAA) were present in the blood serum of subjects with PCS. Taking into account all these considerations, the role of fibrinogen in the relationship between blood group O and PCS myalgia is biologically plausible.

### Blood group O and prevalence of IgG+ antibodies

Hyperimmune plasma from convalescent COVID-19 donors has been a therapeutic option since the beginning of the pandemic. This has opened the option to assess a potential relationship between antibody titers and the ABO system, although the results of these studies have been controversial. Thus, on a sample of 232 plasma donors, Hayes *et al*. [46] observed that blood group O donors had lower titers than the other groups. Prevalences of high titers were 60%, 58%, 65%, and 35% for A, B, AB, and O, respectively. In the same line, Bloch *et al*. [47] analyzed 202 donors and reported that in blood group B there was a significantly higher number of subjects with high titers of neutralizing antibodies when compared with blood group O. Also, Madariaga *et al*. [48], that donors with blood group AB had higher levels of anti-spike and anti-receptor-binding domain (RBD) antibodies than subjects with blood group O.

However, other studies have not confirmed these observations. Žiberna *et al*. [49] analyzed plasma samples from 3185 convalescent donors with 3 serological tests and a neutralizing antibody test. No difference between ABO groups with any of the tests was found. Similar results have been reported in other studies [50-51]. According to these authors, the discrepancies in the results could be explained by the lack of representation of all ethnicities, the heterogeneity of the donors, the severity of the disease, the associated comorbidity, the small sample sizes in the studies carried out at the beginning of the pandemic, and the different sensitivity of the serological tests [50-52].

It is known that high SARS-CoV-2 antibody titers are associated with male gender, older age, and severe acute COVID-19 [47,48], while tobacco use is associated with lower titers [53] and that there is a rapid loss of anti-RBD antibodies in mild COVID-19 [54].

In concordance, IgG+ subjects presented a higher mean age (p=0.03) than those IgG-. The mean age of the whole sample was 45.7±16 years, and all had suffered from mild COVID-19. These two factors (age and mild COVID) could explain the relatively low prevalence of IgG+ antibodies in our population (81%).

There were no significant differences between group O and non-O subjects regarding anti-SARS-CoV-2 IgG. Noteworthy, in IgG+ patients we found a higher number of PCS symptoms (p=0.044), as reported in other studies [55]. Persistently high antibody titers, due to over-activation of the immune system, have been suggested as a risk factor for PCS [55,56].

### Potential implications of the results

Blood O group has been associated with a 4.2-fold increased risk of prevalent PCS compared to non-O subjects, after adjusting for confounders. Group O, lymphocyte and neutrophil counts, and female sex, were the variables that contributed the most in the predictive PCS model. The R^2^ value (0.46) indicates a good fit and a moderate to strong effect size [57], and the AUC of 0.807, an adequate discriminatory ability of the model [58].

Participants with blood group A, with a higher risk of acquiring the disease and developing a worse clinical outcome, according to previous reports, did not show any association with PCS. In contrast, blood group O, that seems to protect against SARS-CoV-2 infection, has shown to be a consistent risk factor for developing PCS, according to our results. Therefore, group O could play a dual role in COVID-19.

The implications of these results deserve a brief comment. Even with the protection of group O subjects from contracting COVID-19, it may imply a high prevalence of PCS in geographic areas with a predominance of this blood group. In the clinical setting, it may be helpful for PCS risk stratification in acute COVID-19. In addition, it can be useful for the diagnosis of a patient with a clinical suspicion of PCS in the post-acute phase. PCS is a challenge for clinicians, as there are no well-defined criteria to define this condition, patient-reported symptoms can be complex, changing and fluctuating, and complementary tests may be of little diagnostic value [59]

This study has some limitations. Firstly, its design allows defining associations, but it does not establish causality. Secondly, it is a single-center study, on a Caucasian population, and the results may not be extrapolated to other populations. The frequencies of the ABO groups in the studied population are a confounding factor since geographical variations constitute potential bias [6]. In our case, the blood group proportions have been very similar to those in Spain (43.3% group A, 43.3% group O, and 13.4% groups B+AB). The main strength in this study is the well-characterized sample and the control group with subjects that came from the same population as the cases, and did not differ from them in any characteristics except for the PCS. Biomarkers have shown slight elevations, without exceeding the upper limit of normality (Figure 2), in a similar way to our previous study [19]. Besides, the use of composite indices of inflammation has increased the ability to detect parameters with values in the upper ranges of their distribution.

## Conclusion

We have found an increased risk of prevalent PCS associated with blood group O. Slightly, albeit significant, raised levels of fibrinogen, CRP, and lymphocyte count, not observed in the other ABO blood groups, have been demonstrated in blood group O.

According to our results, group O could be part of the immunological link between acute COVID-19 and PCS. Further research in large populations is needed to confirm these results and gain more knowledge about PCS pathogenesis.

## Data Availability

All data produced in the present study are available upon reasonable request to the authors

